# Public attention and policy responses to COVID-19 pandemic ^⋆^

**DOI:** 10.1101/2020.06.30.20143420

**Authors:** Cevat Giray Aksoy, Michael Ganslmeier, Panu Poutvaara

## Abstract

Early non-pharmaceutical interventions (NPI) significantly reduced the death toll of the COVID-19 pandemic. Yet, there are vast differences in how quickly governments implemented NPIs. In this paper, we analyze the role of public attention, measured as the share of daily Google searches in a country related to COVID-19, in the timing of the NPI responses. We first show that public attention depends strongly on whether there are cases in own country. We then show that countries with high levels of public attention are more likely to implement non-pharmaceutical interventions, even after controlling for the number of cases and deaths. Finally, we show that the extent to which a government responds to public attention is highly dependent on the country’s institutional quality. The positive effect of public attention on policy implementation is driven entirely by countries with good institutions.

## 1. Introduction

The Coronavirus disease 2019 (COVID-19) was first observed in China in December 2019. Subsequently, the World Health Organization (WHO) declared the outbreak of COVID-19 as a pandemic on 11 March 2020. At the time of the WHO’s declaration, there were 80,955 cases in China with 3126 deaths, and 37,364 confirmed cases, and 1130 deaths in the rest of the world (World Health Organization 2020). As of 29 June 2020, there are more than 10.1 million confirmed cases worldwide and more than 502,000 reported deaths (John Hopkins University 2020). With that said, the infection rates, death toll, and policy responses vary substantially from country to country.

Epidemics are an ultimate stress test for governments. They face the urgent problem of assembling information and mounting effective NPIs (such as travel restrictions, canceling public events, school closures, and so on) against a rapidly spreading virus (Li et al. 2020; Maier and Brockmann 2020). For example, many European countries and the United States have been criticized for being too slow to introduce NPI measures limiting the spread of the COVID-19 virus (Odone et al. 2020; Haffajee and Mello 2020). Slow responses appear to have increased the death toll considerably. According to a recent estimate, the United States could have avoided more than half of reported infections and deaths as of May 3, 2020 by implementing the same control measures just one week earlier (Pei et al. 2020). Singapore, South Korea, and Taiwan, on the other hand, reacted rapidly and contained the pandemic with various NPIs (Lee et al. 2020; Shima et al. 2020; Wang et al. 2020). What explains this difference?

Previous research explaining the introduction of NPIs against the COVID-19 pandemic has studied the role of politicians and political regimes. Adolph et al. (2020) conclude that Republican governors and governors from states in which Trump won a higher share of votes in 2016 were considerably slower to adopt social distancing measures. Frey at al. (2020) find that autocratic regimes imposed more stringent lockdowns as a response to COVID-19 pandemic but were less effective in reducing mobility. Yet, there are wide differences in the speediness of adopting NPIs also between democratic countries. Switzerland implemented the first NPI 3 days after the first confirmed COVID-19 case, Ireland after 8 days, South Korea after 11 days and Singapore after 15 days. The United States waited 34 days, Belgium 35 days, and the United Kingdom 45 days.

National leaders matter for growth (Jones and Olken, 2005), and leaders can make a major difference for better or worse also in disasters (Elinder and Erixson, 2012). Although there is convincing evidence that misinformation and political polarization have played a major role in how the public has responded to COVID-19 in the United States (Allcott et al. 2020; Bursztyn et al. 2020) and in Brazil (Ajzenman et al. 2020), cross-country differences cannot be explained simply by slow responses by populists or right-wing leaders. Pro-European President Macron implemented the first NPI in France 36 days after the first confirmed COVID-19 case, Socialist Prime Minister in Spain after 37 days, and Social Democratic Prime Minister in Sweden after 41 days. Nor does fast reaction by South Korea simply testify of a systematic difference between Western and East Asian countries: Japan waited 36 days before introducing its first NPI.

In this paper, we focus on the public attention part of the equation. Where public attention to COVID-19 is low, governments may face a dilemma when deciding whether to impose NPIs to prevent the spread of the virus. If governments react too little or too late, the death toll may be considerably higher than with earlier intervention (Boorsma and Ferguson 2007; Hatchett et al. 2007; Ferguson et al. 2020; Pei et al. 2020). If early interventions are successful and the epidemic is less lethal than feared, governments risk being accused of overreacting and creating economic and social damage. We conjecture that governments are more likely to take proactive measures in countries with high public attention. Public attention may also provide governments with an opportunity to inform the public on individual measures that would help to reduce the risk of infection (Benzell et al. 2020).

To examine when and to what extent people around the world were alerted to the pandemic, we obtained the daily share of searches related to the COVID-19 of all Google searches in 174 countries from 1 January to 31 March 2020. We then link those searches to the daily data on COVID-19 cases and deaths, as well as governments’ policy responses. Google Trends has previously been used to detect regional disease outbreaks (Ginsberg et al. 2009) and predict economic indicators (Choi and Varian 2012).

Our paper complements recent work on how public attention and anxieties have shaped macroeconomic responses to COVID-19 pandemic. Binder (forthcoming) used Amazon Mechanical Turk to survey respondents in the United States on March 5 and 6, 2020 and found considerable anxiety about the effects of COVID-19 on the economy and personal health and finances. She also found that informing consumers about the Federal Reserve’s recent interest rate cut made consumers less pessimistic. Along the same lines, if consumers are anxious and attentive to the pandemic, NPIs might help to recuperate some of the lost confidence. Fetzer et al. (forthcoming) also used Google searches to analyze the effects of COVID-19 on economic anxiety. They find that both the first confirmed COVID-19 case and the first human-to-human transmission increased searches related to recession, conspiracy, and survivalism. Our paper is complementary. Whereas Fetzer et al. (forthcoming) focus on individual responses to the COVID-19 pandemic, we also analyze to what extent individual attention to the COVID-19 pandemic helps to predict governments’ policy responses. We are also the first to analyze public attention as a share of all Google searches, both nationally and globally. Fetzer et al. (forthcoming) use more restricted Google Trends measure that analyzes how search interest related to certain terms develops over time when the highest attention level is normalized to 100, but without information on what share of all Google searches that corresponds to.

## 2. Data

This section documents our data sources. Descriptive statistics are provided in Table A1 in the appendix.

### 2.1 Google Health Trends API

We measure public attention to COVID-19 using Google Health Trends API for which we applied and obtained access for research purposes. Google Health Trends API allows us to measure the daily share of Google searches related to COVID-19 in each country, as well as globally. We use daily data from 1 January 2020 until 31 March 2020.

In contrast to the publicly available Google Trends database, Google Health Trends API comes with two advantages. First, the actual numbers are not scaled between 0 and 100 but instead, the values represent the share of Google searches that meet our query conditions as a share of all Google searches at a given date in a specific country. Thus, our variables are both more transparent in terms of construction and are continuous. Second, the scaling from 0 to 100 in publicly available Google Trends always depends on the countries and reference terms that are included. Google Health Trends API enables us to achieve cross-country comparability without relying on reference countries.

Furthermore, access to Google Health Trends API enables us to query up to 30 search terms at a time. We download the combined share of Google searches for all queries that included any of the following search terms (connected through the OR-operator): “corona”, “wuhan virus”, “covid”, and “covid-19”. Since Google Health Trends API does not have built-in translation capabilities, we use Google Translate beforehand to translate these four search terms into the national language of each country. Thus, for countries where the first official language is not English, we insert eight search terms (four in English and four in the national language). In addition, we have added “korona” to all search queries as it is a very common name for the disease globally. In countries that do not use the Latin alphabet, we included search terms in both Latin (for terms in English and word korona) and the national writing system.

In terms of country coverage, we start with a global sample of countries. We then drop all countries in which the internet is classified as “Not Free” according to the Internet Freedom Scores provided by Freedom House. When the Internet Freedom scores dataset does not cover a given country in our sample, we use Global Freedom data scores (again from Freedom House) and drop countries that are classified as “Not Free” (Freedom House 2020). Overall, we gathered Google Health Trends API data for 174 countries. In addition to the country level, we also extract Google search volumes for the entire globe.

### 2.2 Cases and death data

We use datasets on COVID-19 cases and deaths from John Hopkins University (John Hopkins University 2020). Starting from these cumulative cases and deaths time series at the country level from the beginning of January until the end of March, we create time dummies that take the value 1 from the date a country has its first case/death, and 0 otherwise. In addition, we merge the global and continental cumulative number of cases/deaths time series to the country-date panel. The continental time series are adjusted by subtracting the cases/deaths of the own country. By doing so, we isolate the continental situation from the country-specific one. Correspondingly, we adjust global time series for each country by subtracting the cases/deaths of the own continent.

### 2.3 Policy data

The Oxford Government Response Tracker (version from 5^th^ of May 2020) provides information on the policy response to the COVID-19 pandemic for 151 countries starting on 1 January 2020 (Hale et al. 2020). Policies are broken down into various categories. For the present project, we examined five domestic social distancing measures, namely *“c1_schoolclosing”, “c2_workplaceclosing”, “c3_cancelpublicevents”, “c5_closepublictransport”* and *“c7_domestictravel”*. Each variable can take value 0 (for no measure in place), 1 (for recommended policy), or 2 (for mandatory policy). We construct a binary variable which is equal to 1 if any of these variables takes value 1 or 2, and 0 if all policy measures are coded 0.

### 2.4 Institutional quality data

We use the World Bank’s Worldwide Governance Indicators (WGI) to measure institutional quality (Kaufmann et al. 2011). The WGI report on six broad dimensions (Voice and Accountability; Political Stability and Absence of Violence; Government Effectiveness; Regulatory Quality; Rule of Law; and Control of Corruption) of governance for over 200 countries and territories over the period 1996-2018. We use the latest available index scores (that is, from 2018) in our analysis (World Bank 2018). The composite measures of governance generated are in units of standard normal distribution, with mean zero, a standard deviation of one, and running from approximately −2.5 to 2.5, with higher values corresponding to better governance. In our analysis, we divide countries in each dimension into those with below-median and those with above-median institutional quality.

## 3. Descriptive patterns

We illustrate a link between COVID-19 cases and deaths, public attention, and NPIs in Fig. 1. In particular, we present the development in the cumulative number of confirmed COVID-19 cases and deaths and public attention, and the timing of first domestic NPI in Singapore, South Korea, the United Kingdom, and the United States. Several patterns are notable. In Singapore and South Korea, public attention increased rapidly after the first domestic case. South Korea imposed domestic restrictions aiming to curtail the epidemic 11 days and Singapore 15 days after the first domestic case was reported, and before a single death. However, public attention remained relatively low for several weeks in the United Kingdom and the United States, and the first domestic restrictions were introduced only after the first deaths. In the United States, the first domestic NPIs were imposed after 34 days, while the UK government waited 45 days before it imposed the first restrictions.

**Fig. 1.**
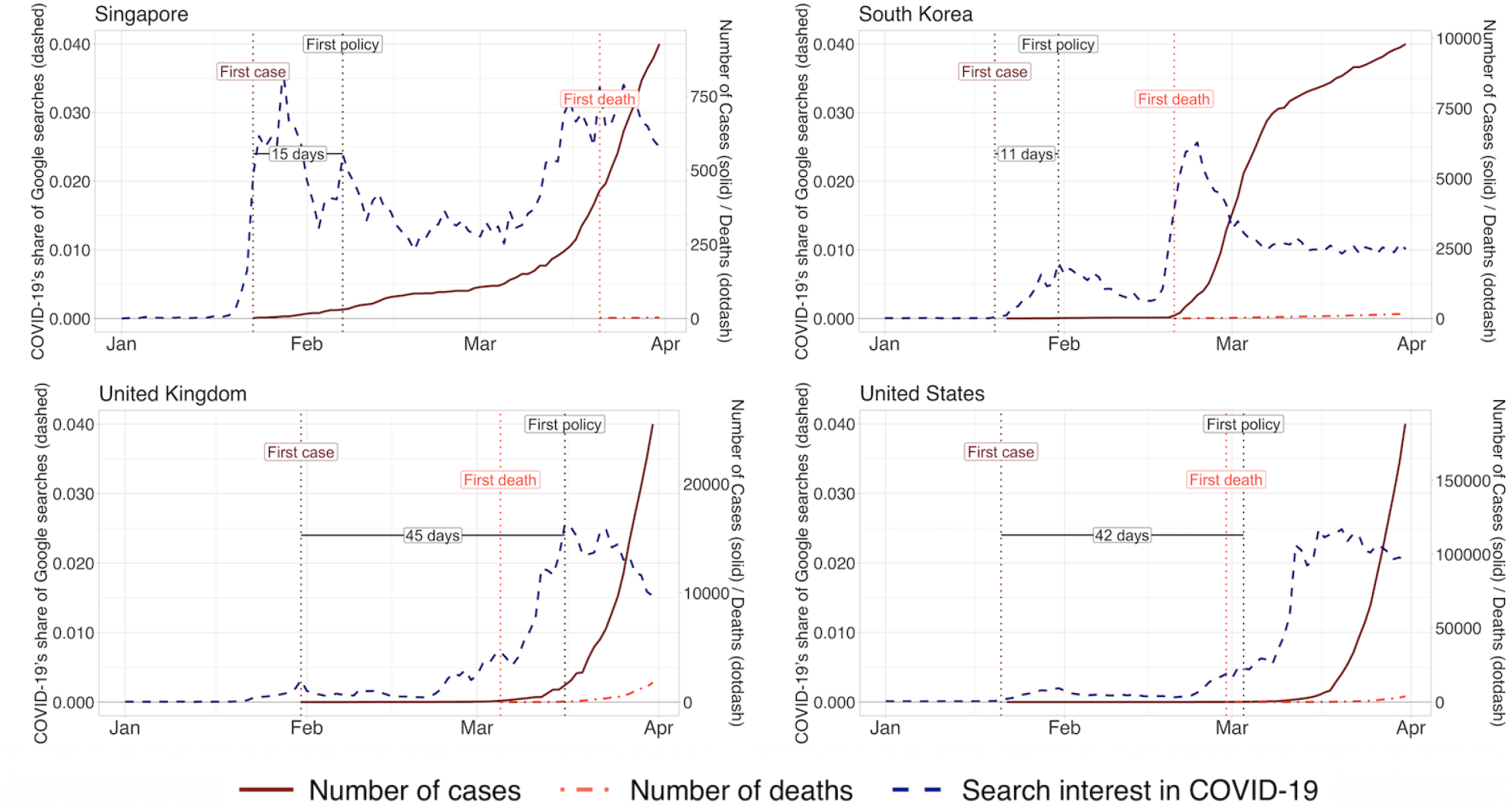
Surge in Google searches predated non-pharmaceutical interventions. Note: The figure depicts daily measures of public attention to COVID-19 measured as a share of daily Google searches (left axis) and the number of COVID-19 cases and deaths (right axis), as well as the dates of the first case, first death, and first policy in Singapore, South Korea, the United Kingdom, and the United States.

Slow reactions were associated with exponential growth a few weeks later. Although Singapore and the United States differ vastly in size, making direct comparisons between the two problematic, populations and land areas in South Korea and the United Kingdom are of a similar order of magnitude. With active testing and NPIs, South Korea had identified 3736 cases on March 1, as compared with only 36 cases in the United Kingdom. Two months later, on May 1, South Korea had 10,780 confirmed cases and the United Kingdom 177,454. The total death toll in South Korea was 250 and in the United Kingdom 27,510 (John Hopkins University 2020).

A potential concern related to Fig. 1 is whether the patterns reflect a cultural difference between Singapore and South Korea on one hand, and the United Kingdom and the United States on the other hand. As shown in Fig. A1 in the appendix, a similar pattern as in Singapore and South Korea prevails in those European countries in which public attention increased fast. Denmark introduced the first NPI 8 days after the first case, Ireland after 9 days, Portugal after 10 days and Switzerland after only 3 days. Slow response when public attention remained weak was not specific to the United Kingdom and the United States. Belgium, France, Spain, and Sweden also had initially a low level of public attention, and governments in those countries waited more than a month before the first NPI was introduced (Fig. A2 in the appendix).

Our examples suggest that both policy-makers and public attention are important for the timing of NPIs. Although France and Spain introduced the first NPI five weeks after the first case was confirmed, the implementation took place when public attention was still relatively low, compared with levels at which first NPI was introduced in most countries. In Sweden and the United Kingdom, instead, policymakers initially appeared reluctant to impose NPIs, and introduced restrictions only after public attention and related public pressure had reached a high level.

The descriptive patterns reported in Fig. 1 suggest a link between COVID-19 cases and public attention. Next, we plot this link in a global sample. In Fig. 2, we show how global attention to the COVID-19 pandemic and the global number of COVID-19 cases and deaths developed on a daily level. Global cases and deaths remained at a relatively low overall level until mid-March, from which onwards the exponential growth path is clearly visible on a global scale.

**Fig. 2.**
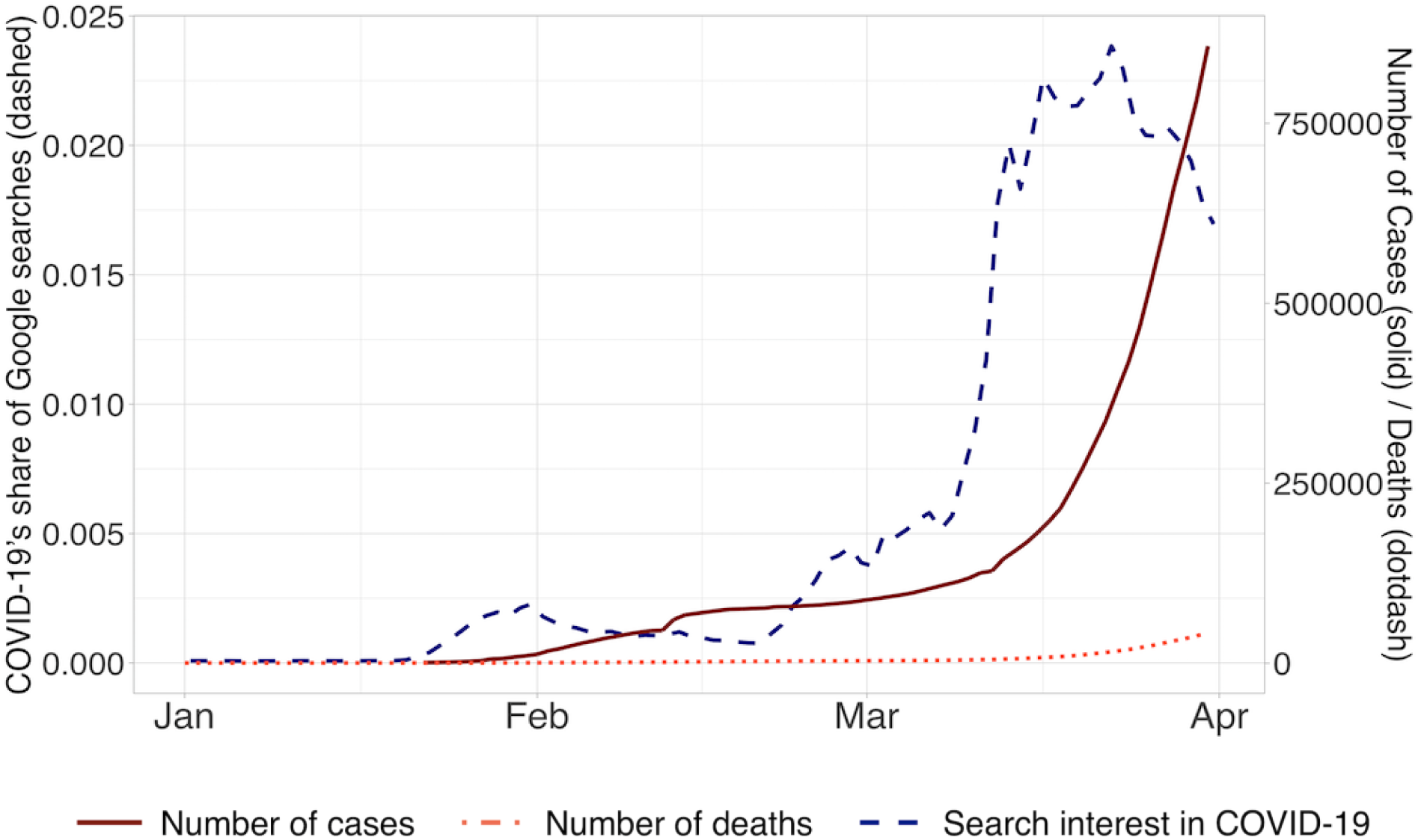
Global Google searches related to COVID-19 pandemic increased dramatically in March. Notes: The figure depicts the development of public attention to COVID-19 pandemic at the global level (left axis) and the global number of cases and deaths (right axis) for each day from 1 January to 31 March 2020.

Figs 3A, 3B, and 3C provide snapshots on attention around the world at the end of January, February, and March. Comparing these figures with the spread of cases (Fig. A3) and deaths (Fig. A4) in the appendix suggests a clear pattern: Attention is to a large extent local. When the virus was mostly limited to Asia, attention in Europe and the Americas remained low.

**Fig. 3.**
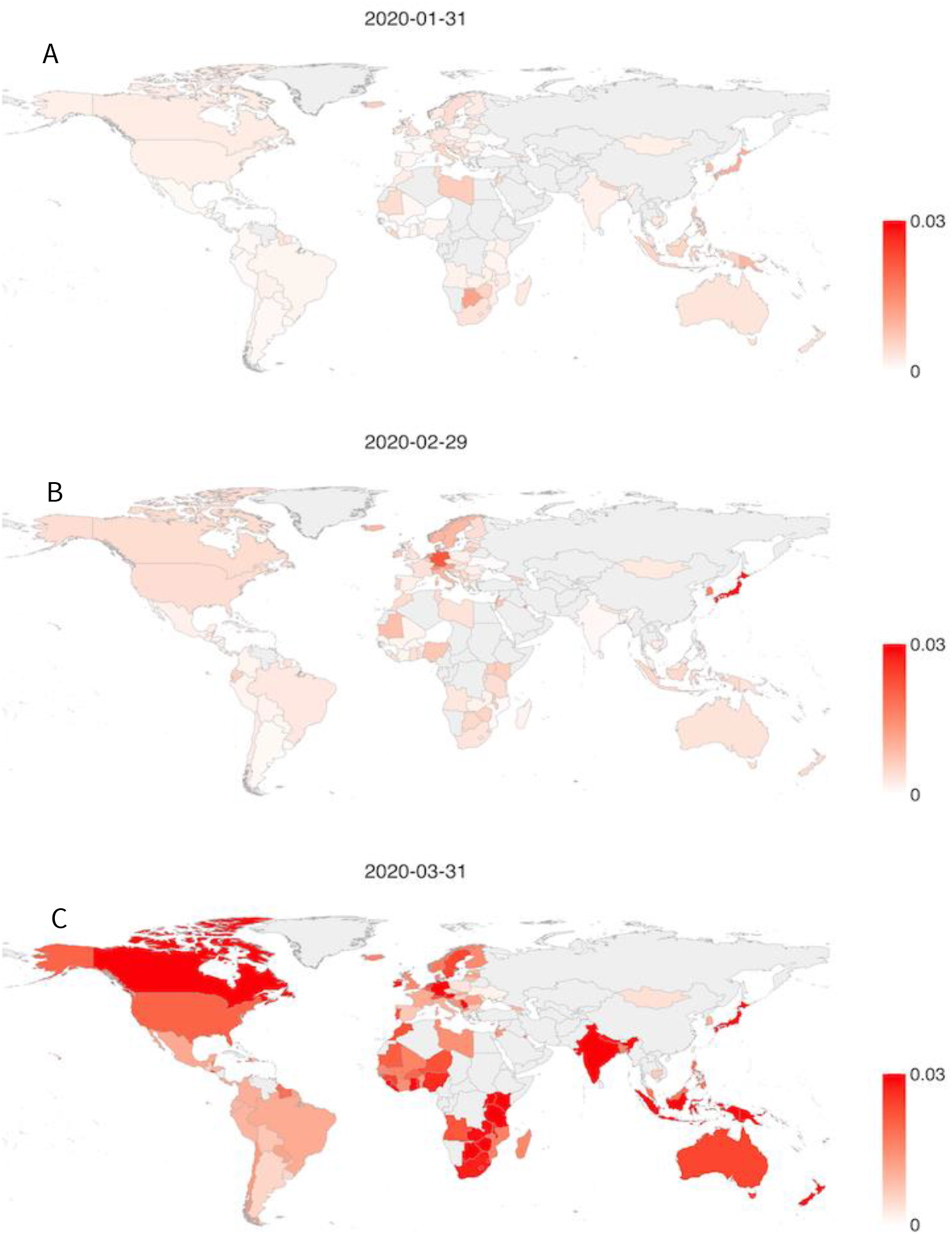
Public attention spread globally following the epidemic. Notes: Panels A, B, and C depict the share of Google searches related to COVID-19 in all countries on 31 January (A), 29 February (B), and 31 March (C). Darker red corresponds to higher public attention.

## 4 Econometric analysis

### 4.1 Event study

We estimate the relationship between the first confirmed case in the own country and changes in public attention to COVID-19 using an event study methodology. We proxy public attention by Google searches. The main variable of interest (that is, event) is an indicator variable equal to one in the countries and days when the first COVID-19 case is officially observed. The sample consists of 78 countries that ever-adopted non-pharmaceutical policy between 1 January and 31 March 2020. We control for (log) own continent cases excluding own country plus one, and (log) global cases excluding own continent plus one. We also include country and date fixed effects. The former absorbs time-invariant variation in the outcome variable caused by factors that vary across countries while the latter eliminates time-varying shocks that affect all countries simultaneously. In the presence of country and year dummies, the effect is identified from sharp within-country changes in outcome coincident with variation in the timing of the first confirmed COVID-19 case. Standard errors are clustered at the country level.

We present event study estimates of the relationship between the public attention and the first case in own country as Fig. 4. We exclude early weeks as in Adukia et al. (2020) and Aksoy et al. (2020) and leave out the week just prior to policy adoption since there may have been discussion on NPIs already in the days before it.

**Fig. 4.**
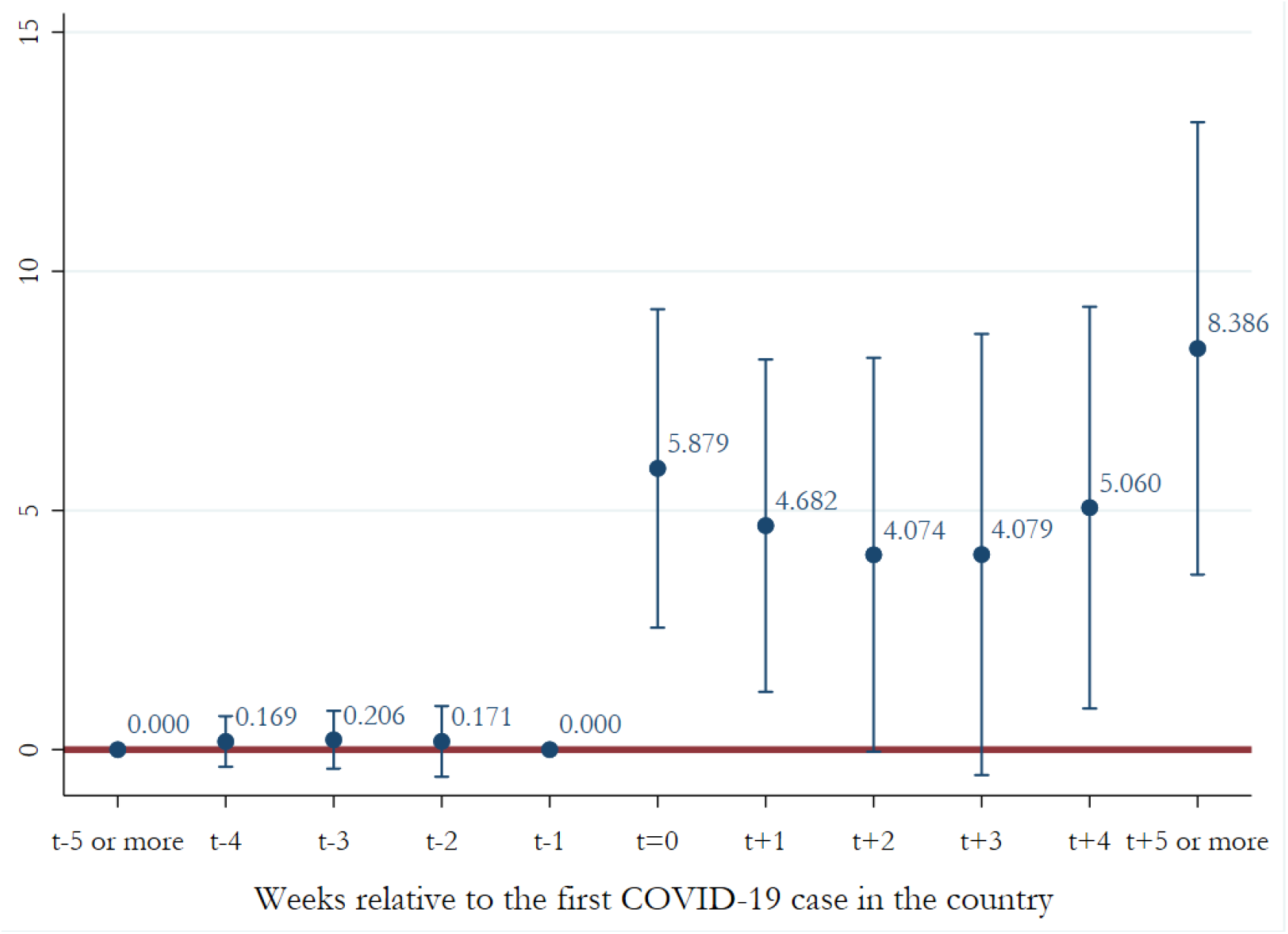
Public attention to COVID-19 increases strongly after first domestic case is confirmed. Notes: This figure shows event-study estimates of the relationship between the first confirmed case in the own country and changes in public attention to COVID-19. The main variable of interest is an indicator variable equal to one in the countries and days when the first COVID-19 case is officially observed. It is based on OLS specification which includes controls for (log) own continent cases excluding own country plus one, (log) global cases excluding own continent plus one, country dummies, and date dummies. In the presence of country and year dummies, the effect is identified from sharp within-country changes in outcome coincident with variation in the timing of the first confirmed COVID-19 case. Standard errors are clustered at the country level. The sample consists of countries that ever-adopted non-pharmaceutical policy between 1 January and 31 March 2020. Sources: Google Health API and Johns Hopkins Coronavirus Resource Center.

We find that the first case in own country led to substantial increases in public attention toward COVID-19 related topics in the following weeks. The pre-trend coefficients are close to 0 and statistically insignificant, satisfying the ‘no pre-trends’ assumption required for identification in event studies. Fig. 4 also indicates that public attention to the COVID-19 lasts several weeks: the coefficients on the post-period indicators are positive, sizable, and mostly statistically significant.

### 4.2 The effect of public attention on NPIs

Finally, we link public attention to NPIs in Table 1. We start by analyzing 78 countries with at least partially-free internet and that ever-adopted non-pharmaceutical policy between 1 January and 31 March 2020 and that satisfied the criteria to be included in the event study analysis. We then analyze the role of institutional quality. Countries with high institutional quality presumably are more able to mount effective responses to the pandemic. If they are more prone to listening to the public, we might expect the effects we identify to be strongest when the institutional quality is high, other things equal. To explore this hypothesis, we use the World Bank’s Worldwide Governance Indicators.

**Table 1:** Public attention to COVID-19 significantly increases likelihood of policy intervention.

| Sample | (1)<br>Coefficient on the share<br>of COVID-19-related<br>Google searches (average<br>within the previous 10<br>days) (standard error) | (2)<br>Coefficient on (log)<br>own country cumulative<br>cases + 1<br>(standard error) | (3)<br>Coefficient on (log)<br>own continent cases + 1<br>(exc. own country)<br>(standard error) |
| --- | --- | --- | --- |
| Full-sample | 0.060*** [0.061]<br>(0.022) | 0.044*** [0.115]<br>(0.010) | 0.028*** [0.132]<br>(0.009) |
| N | 6318 | 6318 | 6318 |
| Below median control of corruption | 0.032 [0.020]<br>(0.032) | 0.044* [0.081]<br>(0.023) | 0.035** [0.159]<br>(0.017) |
| Below median government effectiveness | 0.023 [0.021]<br>(0.034) | 0.048* [0.083]<br>(0.024) | 0.043* [0.145]<br>(0.018) |
| Below median political stability | 0.000 [0.001]<br>(0.026) | 0.040** [0.086]<br>(0.016) | 0.031** [0.196]<br>(0.014) |
| Below median regulatory quality | 0.030 [0.027]<br>(0.031) | 0.057** [0.105]<br>(0.023) | 0.039** [0.177]<br>(0.015) |
| Below median rule of law | 0.029 [0.026]<br>(0.032) | 0.054** [0.098]<br>(0.023) | 0.044** [0.201]<br>(0.018) |
| Below median accountability | 0.047 [0.045]<br>(0.036) | 0.068*** [0.130]<br>(0.025) | 0.048*** [0.226]<br>(0.017) |
| N | 3159 | 3159 | 3159 |
| Above median control of corruption | 0.080**<sup>A</sup> [0.089]<br>(0.035) | 0.054*** [0.162]<br>(0.015) | 0.018 [0.084]<br>(0.012) |
| Above median government effectiveness | 0.078**<sup>A</sup> [0.087]<br>(0.035) | 0.050*** [0.130]<br>(0.016) | 0.014 [0.069]<br>(0.011) |
| Above median political stability | 0.107***<sup>A</sup> [0.116]<br>(0.034) | 0.044** [0.152]<br>(0.017) | 0.015 [0.065]<br>(0.013) |
| Above median regulatory quality | 0.087**<sup>A</sup> [0.095]<br>(0.035) | 0.039** [0.119]<br>(0.016) | 0.016 [0.074]<br>(0.011) |
| Above median rule of law | 0.083**<sup>A</sup> [0.092]<br>(0.035) | 0.041** [0.124]<br>(0.016) | 0.013 [0.059]<br>(0.011) |
| Above median accountability | 0.074**<sup>A</sup> [0.080]<br>(0.033) | 0.041** [0.123]<br>(0.016) | 0.017 [0.078]<br>(0.011) |
| N | 3,159 | 3,159 | 3,159 |

Notes: \* significant at 10%; \*\* significant at 5%; \*\*\* significant at 1%. OLS regressions. Each model also controls for country fixed effects, date fixed effects, (log) own country cumulative cases plus one, (log) own continent deaths plus one, (log) GDP per capita, (log) urbanization rate. Robust standard errors are clustered at the country level. Brackets report point estimates for one standard deviation increase in Google searches (lagged ten days). <sup>A</sup> indicates statistically significant differences between the pair estimates. The sample consists of 78 countries that ever-adopted non-pharmaceutical policy between 1 January and 31 March 2020. Sources: Google Health API, Johns Hopkins Coronavirus Resource Center, the Oxford COVID-19 Government Response Tracker, and World Bank Governance Indicators.

We estimate OLS regressions to investigate the relationship between public attention and the likelihood of policy intervention. In each model, we control for (log) own country cumulative cases plus one, (log) own continent deaths plus one, (log) GDP per capita, (log) urbanization rate. We also include a full set of country fixed and date fixed effects. Robust standard errors are clustered at the country level.

The first panel of Table 1 shows that a one standard deviation increase in the share of COVID-19-related Google searches (average within the previous 10 days) is associated with 6.1 percentage points higher likelihood of NPI being in place on a given day. When we compare standardized coefficients, the effect of Google searches is half as strong as the effect of cases in the own country, and also half as strong as the effect of cases in the other countries on the own continent.

We next analyze how the country characteristics matter when it comes to taking public attention into account. In the second and third panel of Table 1, we use the same specification as in the first panel when we split the sample by six measures of institutional quality. To do so, we use the World Bank’s Worldwide Governance Indicators (Kaufmann et al. 2011), which provide institutional quality scores for a global sample of countries. The second and third panels show that the effect of public attention is driven by countries with high institutional quality. Instead, there is no statistically significant association between public attention and NPI in countries with poor institutional quality. This is in line with the recent studies finding that weak and fragmented governments fail to take appropriate policy actions in the wake of financial crises and economic downturns (Caselli and Tesei 2011; Cox and Weingast 2018).

To obtain an estimate on how large the effect of public attention is in countries with high institutional quality, we use standardized coefficients. The standardized coefficient of Google searches on the likelihood of NPI varies between 55 and 80% of the standardized coefficient of cases in the own country across the six measures of institutional quality.

An important caveat is that our findings should be interpreted as establishing correlational patterns, rather than as causal estimates on the effects of attention on the probability of introducing an NPI. This is unavoidable when analyzing public policies towards a pandemic threat: randomly assigning public attention to estimate its effects on policies is infeasible and would be ethically questionable and unlikely to obtain IRB approval even if having resources to implement such randomization. Nonetheless, we find it unlikely that our findings would simply reflect a potential correlational pattern that citizens who care more about public health also elect politicians who respond more strongly to health risks like a pandemic. The reason is that we already include country fixed effects that capture also differences between politicians; none of the countries in our analysis experienced a change in the government during the time period we analyze. Furthermore, one could expect that politicians who care more about public health would react already to global cases, or at least to cases in the own continent.

## 5. Conclusion

In this study, we showed that there is a strong link between public attention and the timing of NPIs in countries with high institutional quality. Countries with high institutional quality in which public attention increased fast after the first case, like Denmark, Ireland, South Korea, and Switzerland, introduced NPIs soon after the first case was confirmed. Countries with high institutional quality but low public attention, like France, Spain, Sweden, the United Kingdom, and the United States, waited more than a month before introducing the first NPI. These slow responses go together with very high subsequent death tolls.

Taken together, our findings suggest that in countries with good institutions, high public attention could help to save lives by encouraging governments to respond swiftly. Clearly, these are not alone sufficient conditions. If the government postpones NPIs, despite the strong public attention, the death toll is likely to be much higher. Even though our findings should be interpreted as establishing correlational patterns, they could help in saving lives in eventual subsequent pandemic waves. Identifying how strongly the public responds to pandemic threat helps policymakers in their planning. Strong public attention appears both to give politicians a cover to act fast and may help to persuade even politicians who are initially unwilling to impose NPIs.

NPIs are also important for economic reasons. For example, Binder (forthcoming) shows that informing consumers about the Federal Reserve’s recent interest rate cut made consumers less pessimistic. This raises an intriguing question: could also NPIs help to return confidence in the economy? In a recent paper, Andersen et al. (2020) compare drop in consumer spending in Denmark that introduced drastic NPIs and Sweden that did not. Consumer spending dropped 25% in Sweden and 29% in Denmark, suggesting that most of the decrease in consumer spending was not because of lockdown. Instead, fear of the pandemic is enough to paralyze the economy, even without NPIs. If NPIs succeed in suppressing the pandemic, they can be expected to help also the economy to recover. An important challenge for future crises is the pattern that public attention tends to be local and responds mostly to cases in the own country. Earlier responses of public attention to a global pandemic could both promote international solidarity and save lives domestically if it would encourage politicians to respond proactively.

## Data Availability

We used publicly available data and data from Google Health Trends API for which we applied and obtained access for research purposes. Publicly available data consists of:
(1) Hale, T., Webster, S., Petherick, A., Phillips, T. & Kira, B. (2020). Oxford COVID-19 Government Response Tracker, Blavatnik School of Government. Data use policy: Creative Commons Attribution CC BY standard. Available at: https://www.bsg.ox.ac.uk/research/research-projects/coronavirus-government-response-tracker
(2) Johns Hopkins University, COVID-19 Data Repository by the Center for Systems Science and Engineering (CSSE) (2020); Cases: https://raw.githubusercontent.com/CSSEGISandData/COVID-19/master/csse_covid_19_data/csse_covid_19_time_series/time_series_covid19_confirmed_US.csv; Deaths: https://raw.githubusercontent.com/CSSEGISandData/COVID-19/master/csse_covid_19_data/csse_covid_19_time_series/time_series_covid19_deaths_global.csv
(3) World Bank. Worldwide Governance Indicators (WGI) Project. The World Bank, Washington DC, USA (2018). Available at: https://info.worldbank.org/governance/wgi/

## 1. APPENDIX

**Fig. A1.**
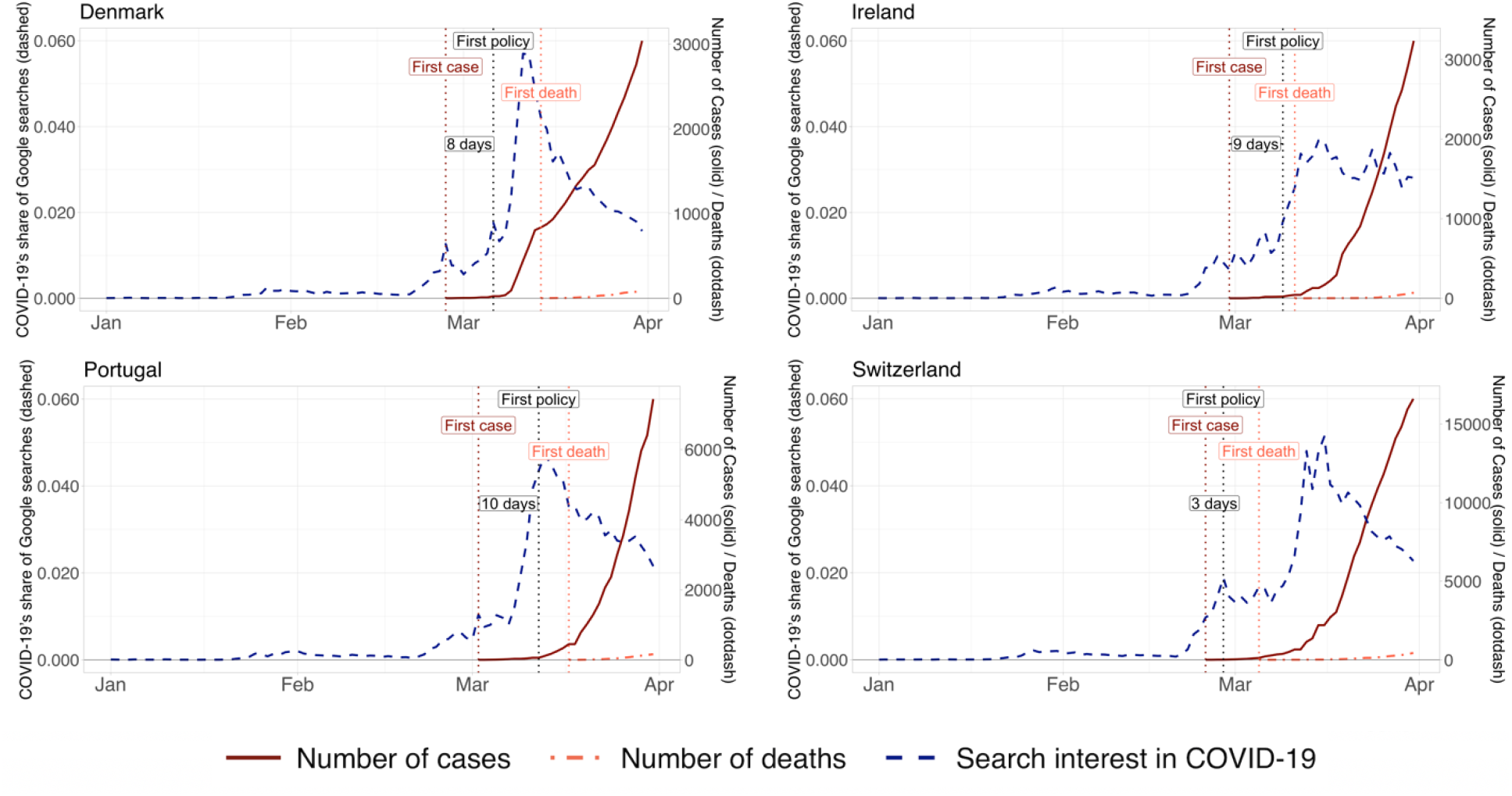
Early increase in Google searches followed by fast non-pharmaceutical interventions.

**Fig. A2.**
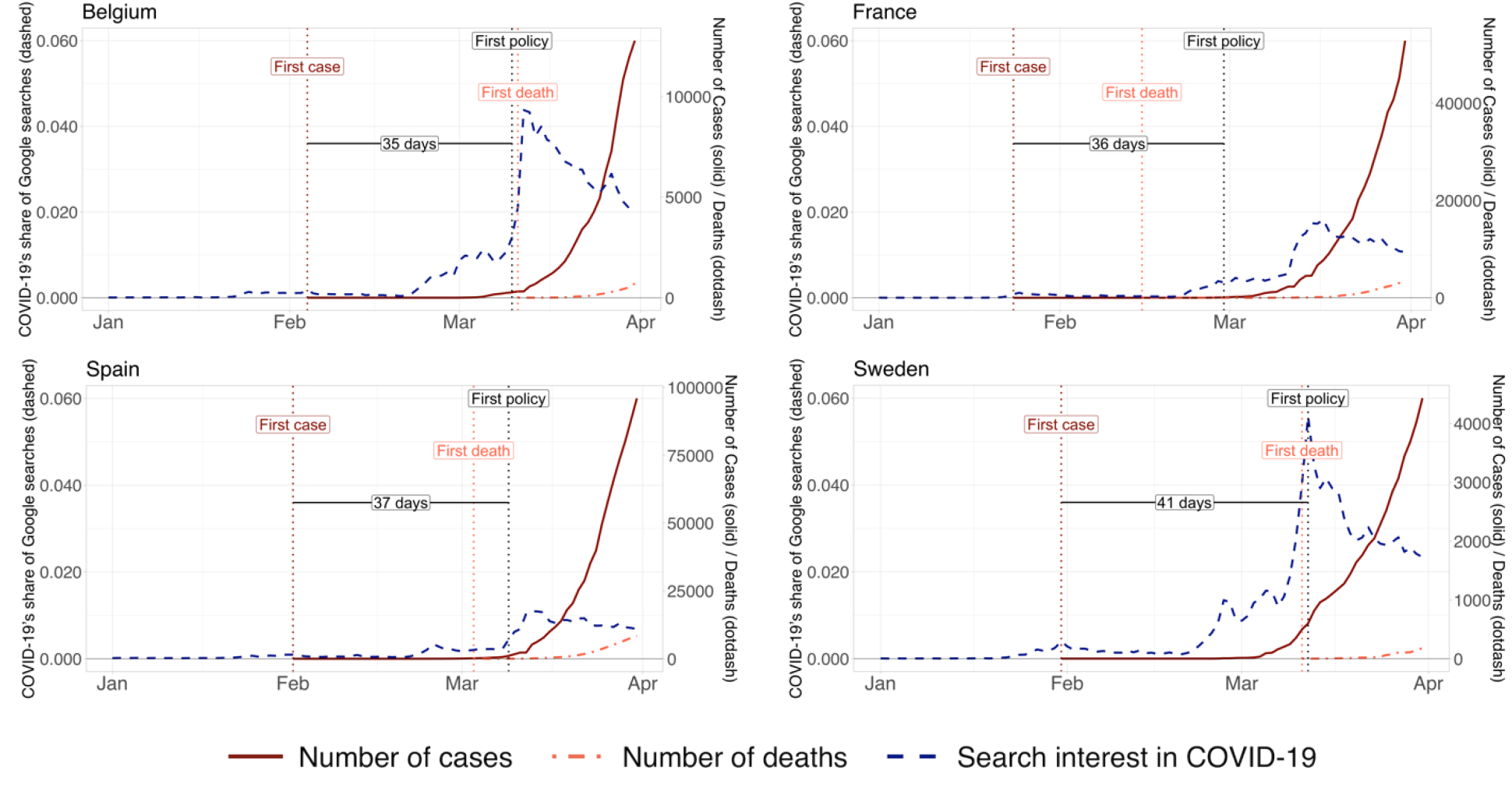
Late increase in Google searches followed by late non-pharmaceutical interventions.

**Fig A3.**
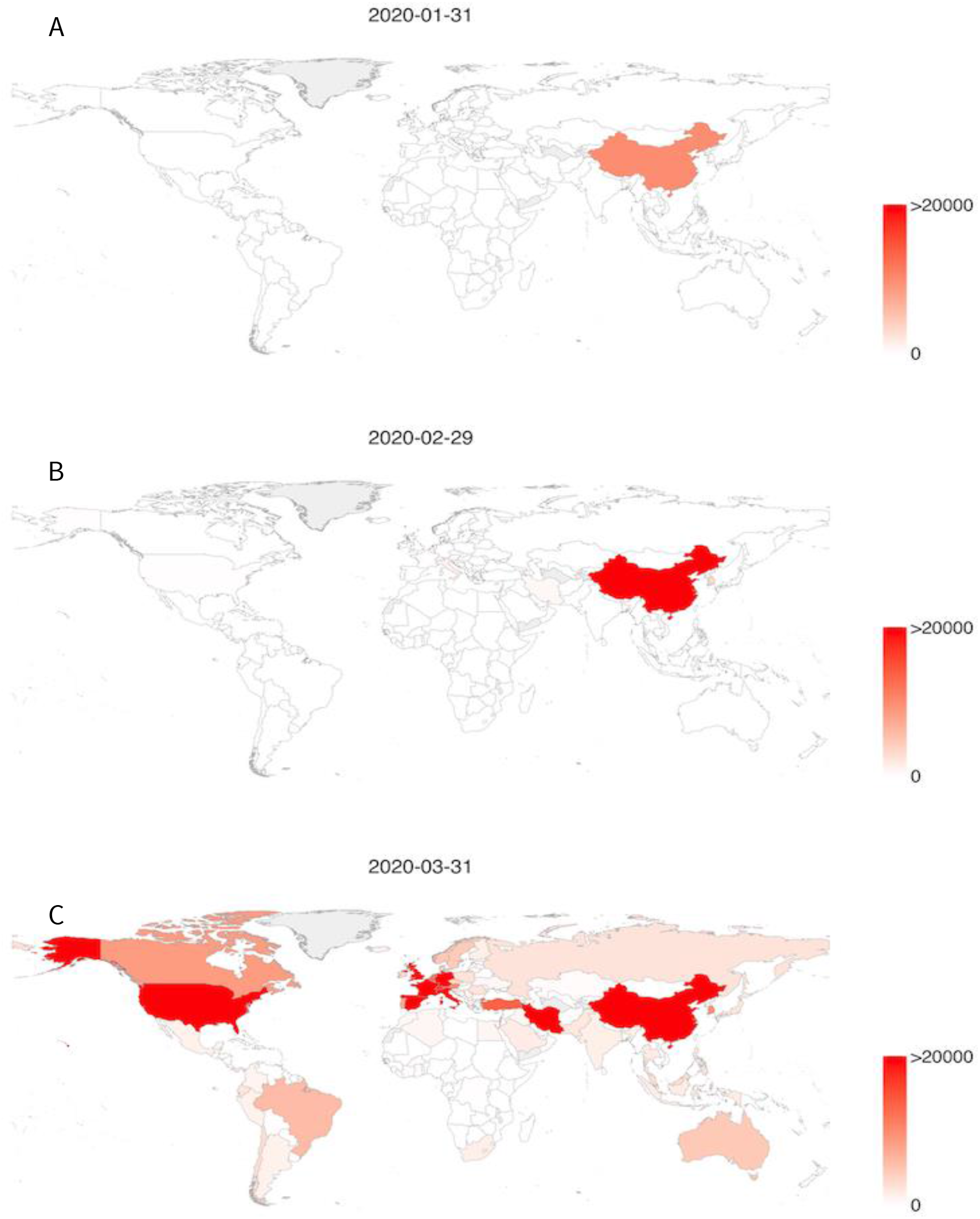
Europe and the United States became COVID-19 epicentres in March. Notes: Panels A, B, and C depict the number of reported COVID-19 cases in all countries on 31 January (A), 29 February (B), and 31 March (C). Darker red corresponds to higher number of cases.

**Fig. A4.**
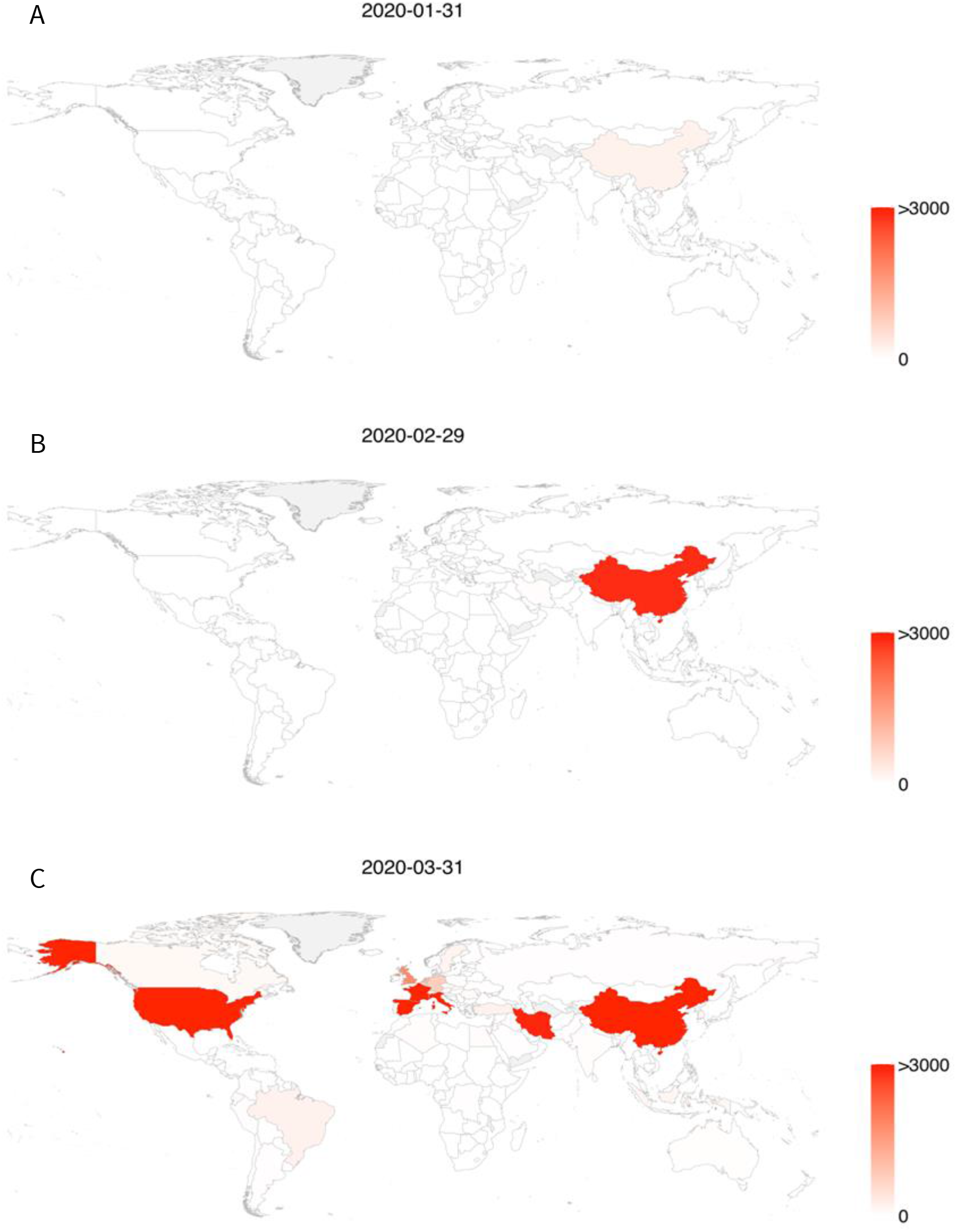
COVID-19 deaths rapidly spread globally in March. Notes: Panels A, B, and C depict the number of reported COVID-19 deaths in all countries on 31 January (A), 29 February (B), and 31 March (C). Darker red corresponds to a higher number of deaths.

**Table A1.** Descriptive Statistics.

| **Variable** | **Source** | **Mean** | **Median** | **Min** | **Max** | **SD** |
| --- | --- | --- | --- | --- | --- | --- |
| Share of covid-19-related Google searches in a country on a given day | Google | 0.007 | 0.001 | 0.000 | 0.333 | 0.013 |
| Total no of COVID-19 cases in a country on a given day | John Hopkins Uni. | 814 | 0 | 0 | 188172 | 6886 |
| Total no of COVID-19 deaths in a country on a given day | John Hopkins Uni. | 33 | 0 | 0 | 12428 | 368 |
| Policy indicator (dummy) for domestic non-pharmaceutical policy measures in a country on a given day | Blavatnik School of Government (Oxford) | 0.27 | 0.00 | 0 | 1 | 0.44 |
| Control of Corruption | World Bank | -0.002 | -0.195 | -1.800 | 2.212 | 1.002 |
| Government Effectiveness | World Bank | 0.028 | -0.043 | -2.449 | 2.231 | 0.995 |
| Political Stability | World Bank | -0.073 | 0.032 | -2.747 | 1.939 | 1.013 |
| Regulatory Quality | World Bank | 0.043 | -0.048 | -2.334 | 2.206 | 0.993 |
| Rule of Law | World Bank | -0.008 | -0.148 | -2.339 | 2.046 | 1.006 |
| Voice and Accountability | World Bank | -0.021 | 0.040 | -2.176 | 1.733 | 0.977 |

